# Blood Pressure Severity Modifies the Association Between Atrial Cardiopathy and Stroke Mortality

**DOI:** 10.64898/2026.08.04.26359746

**Authors:** Asem M. Mohsen, Moustafa Elnewishy, Patrick Cheon, Parag A. Chevli, Brian C. Boursiquot, Richard Kazibwe, Prashant D. Bhave, Elsayed Z. Soliman

**Affiliations:** Epidemiological Cardiology Research Center, Department of Cardiovascular Medicine, Wake Forest University School of Medicine, Winston Salem, NC; Wake Forest University School of Medicine, Winston Salem, NC; Division of Cardiology, Department of Medicine, Columbia University Irving Medical Center, New York, NY; Department of Cardiovascular Medicine, Wake Forest University School of Medicine, Winston Salem, NC; Department of Internal Medicine, Wake Forest University School of Medicine, Winston Salem, NC

**Author notes:** **Conflict of Interest**: BCB has consulted for Ambience Healthcare. The other authors have no relevant financial or non-financial interests to disclose. **Corresponding Author**: Elsayed Z. Soliman MD, MSc, MS, Epidemiological Cardiology Research Center (EPICARE) Wake Forest School of Medicine, Medical Center Blvd, Winston Salem, NC 27157.

**Keywords:** Atrial cardiopathy, P-wave indices, Hypertension, Electrocardiography, Stroke

## Abstract

**Background:** Electrocardiographic (ECG) markers of atrial cardiopathy (AC) are associated with stroke mortality, but whether this association is modified by blood pressure (BP) is unknown.

**Methods:** We analyzed 7,191 adults free of cardiovascular disease from the Third National Health and Nutrition Examination Survey who underwent baseline ECG. AC was defined by three ECG markers: prolonged P-wave duration (≥120 ms), abnormal P-wave axis (<0° or >75°), and deep terminal negativity of the P wave in V1 (<−100 μV). AC burden (per additional AC marker) and AC presence (≥1 vs. 0 markers) were examined in relation to stroke mortality using Cox proportional hazards models. Participants were stratified by BP as normal/elevated (<130/80 mmHg), stage 1–2 hypertension (130–159/80–99 mmHg), or severe hypertension (≥160/100 mmHg). Interaction by BP category was assessed.

**Results:** During a median follow-up of 13.8 years, 183 stroke deaths occurred. In multivariable adjusted model, AC burden was associated with a 41% higher risk of stroke mortality (HR (95%CI): 1.41 (1.13–1.77)). This association was significantly modified by BP (interaction *P*=0.003). The HRs (95% CIs) per additional AC marker were 0.88 (0.52–1.49), 1.39 (1.03–1.88), and 2.94 (1.82–4.75) for normal/elevated BP, stage 1–2 hypertension, and severe hypertension, respectively. A similar pattern of associations was observed for AC presence, although the interaction with BP was not statistically significant.

**Conclusions:** ECG-defined AC burden was independently associated with stroke mortality, with substantially stronger associations among individuals with severe hypertension, supporting BP as an important modifier of its prognostic significance.

## Introduction

Embolic stroke of undetermined source (ESUS) and related cryptogenic stroke phenotypes account for up to one-third of ischemic strokes, yet their underlying mechanisms remain incompletely understood. ^1^ Atrial cardiopathy (AC), a spectrum of structural, electrical, and functional atrial abnormalities, has emerged as a potential substrate for thromboembolism independent of overt atrial fibrillation (AF).^2–4^

Although no standardized clinical definition or diagnostic criteria currently exist for AC, electrocardiographic (ECG) P-wave indices—including prolonged P-wave duration, abnormal P-wave axis, and deep terminal negativity of the P wave in lead V1 (DTNPV1)—are established markers of atrial electrical remodeling that may reflect the underlying atrial substrate. These noninvasive, inexpensive, and widely available measures offer scalable tools for population-level risk stratification.^5,6^ Previous studies have shown that these ECG markers are individually associated with AF, stroke, and mortality,^7–9^ and that a greater cumulative burden of AC markers confers progressively higher risk in a dose-dependent manner.^10^ However, the clinical significance of these markers may depend on the vascular milieu in which they occur rather than being uniform across populations.

Hypertension is a major determinant of cerebrovascular disease and promotes stroke through multiple mechanisms, including small-vessel disease, large-artery atherosclerosis, and cardiac remodeling.^11,12^ Because hypertension is also closely linked to atrial structural and electrical remodeling, it may modify the relationship between AC and stroke outcomes. Whether elevated BP amplifies the prognostic significance of ECG-defined AC or whether alternative hypertensive stroke mechanisms predominate remains uncertain. We therefore investigated whether BP modifies the association between ECG-defined AC and stroke mortality in a cohort of U.S. adults. We evaluated both AC burden (0–3 ECG markers) and AC presence (≥1 vs. 0 markers) overall and across BP strata, hypothesizing that the association between AC and stroke mortality would differ according to BP interesting.

## Methods

### Study Population

We analyzed data from the Third National Health and Nutrition Examination Survey (NHANES III), a nationally representative survey of the U.S. civilian population conducted by the National Center for Health Statistics (NCHS) between 1988 and 1994. All participants provided written informed consent, and the study protocol was approved by the NCHS Institutional Review Board. Detailed descriptions of the survey design have been published previously.^13,14^

Because electrocardiograms (ECGs) were obtained only in participants aged ≥40 years, the present analysis was restricted to this population. We excluded participants with a history of cardiovascular disease (myocardial infarction, heart failure, or stroke), non-sinus rhythm, use of antiarrhythmic medications, electrocardiographic evidence of probable or possible myocardial infarction according to Minnesota codes,^15^ invalid blood pressure measurements, or incomplete follow-up. The final analytic cohort comprised 7,191 participants.

### ECG Assessment of Atrial Cardiopathy

Standard 12-lead ECGs were obtained during mobile examination center visits using a Marquette MAC-12 electrocardiograph (Marquette Medical Systems, Milwaukee, WI). ECGs were transmitted to the Epidemiological Cardiology Research Center (EPICARE), Wake Forest University School of Medicine, where they underwent centralized quality control and automated analysis using the GE 12-SL 2001 program.

Consistent with previous studies,^16–19^ atrial cardiopathy (AC) was defined using three ECG markers: (1) prolonged P-wave duration (≥120 ms in lead II), (2) abnormal P-wave axis (<0° or >75°), and (3) deep terminal negativity of the P wave in lead V1 (DTNPV1 <−100 μV). AC was evaluated as both a burden score (0–3 abnormal markers) and a binary variable (presence of ≥1 vs. 0 markers).

### Blood Pressure Classification

Participants were categorized according to baseline blood pressure (BP) as: (1) normal/elevated BP (systolic BP <130 mmHg and diastolic BP <80 mmHg); (2) stage 1–2 hypertension (systolic BP 130–159 mmHg or diastolic BP 80–99 mmHg); or (3) severe hypertension (systolic BP ≥160 mmHg or diastolic BP ≥100 mmHg). These categories were prespecified to represent increasing hypertensive vascular burden.

### Outcome Ascertainment

The primary outcome was stroke mortality. Mortality status was determined through linkage with the National Death Index using a validated probabilistic matching algorithm. Stroke deaths were identified using International Classification of Diseases, Tenth Revision (ICD-10) codes. Follow-up time was calculated from the baseline examination until stroke death or censoring.

### Covariates

Covariates were selected a priori based on established associations with stroke and cardiovascular disease. Age, sex, race and /ethnicity, educational attainment, smoking status, medication use, and medical history were obtained by standardized interview. Body mass index (BMI) was calculated from measured height and weight. Diabetes mellitus was defined as fasting plasma glucose ≥126 mg/dL or use of glucose-lowering medication. Total cholesterol and high-density lipoprotein (HDL) cholesterol were measured using standardized laboratory methods.

### Statistical Analysis

Baseline characteristics were compared across BP categories using one-way analysis of variance for continuous variables and chi-square tests for categorical variables. Continuous variables are presented as mean ± standard deviation and categorical variables as counts (percentages).

Stroke mortality incidence rates were calculated using person-time methods and expressed per 1,000 person-years overall, by AC burden, by BP category, and across joint strata of AC burden and BP.

Associations between AC and stroke mortality were evaluated using Cox proportional hazards regression. AC was modeled as (1) a continuous variable (per one additional ECG marker) and (2) a binary variable (≥1 vs. 0 markers). Two sequential multivariable models were constructed. Model 1 adjusted for age, sex, race and/ethnicity, and education. Model 2 additionally adjusted for diabetes mellitus, smoking status, BMI, total cholesterol, HDL cholesterol, and antihypertensive medication use. Overall analyses additionally included BP category in Model 2.

Effect modification by BP was evaluated by conducting BP-stratified Cox regression analyses and testing multiplicative interaction terms between AC and BP category using Wald statistics. Interaction analyses were performed separately for AC burden and AC presence.

Kaplan–Meier curves were generated to illustrate cumulative stroke mortality according to AC status and BP category using the product-limit method.

All statistical tests were two-sided, and a *P* value <0.05 was considered statistically significant. Analyses were performed without application of NHANES sampling weights because the primary objective was estimation of associations rather than population prevalence; variables incorporated into the sampling design were included in multivariable models to reduce potential bias.

## Results

Among 7,191 participants (mean age, 58.9 ± 13.3 years; 53.4% women), 2,835 (39.4%) had normal/elevated blood pressure (BP), 3,609 (50.2%) had stage 1–2 hypertension, and 747 (10.4%) had severe hypertension. Compared with participants with normal/elevated BP, those with higher BP were older, had fewer years of education, higher body mass index, a greater prevalence of diabetes mellitus, higher total cholesterol levels, and were more likely to use antihypertensive medications. The prevalence of ECG-defined atrial cardiopathy also increased across BP categories, with prolonged P-wave duration, abnormal PTFV1, greater atrial cardiopathy burden, and the presence of ≥1 atrial cardiopathy marker becoming progressively more common with increasing BP severity (**Table 1**)

**Table 1.** Baseline Characteristics.

| Characteristic* | Overall | Normal/elevated BP | Stage 1–2 HTN | Severe HTN | p-value |
| --- | --- | --- | --- | --- | --- |
| N | 7,191 | 2,835 | 3,609 | 747 |  |
| Age, years | 58.9 ± 13.3 | 54.5 ± 12.1 | 60.7 ± 13.2 | 67.3 ± 12.7 | <0.001 |
| Female sex | 3839 (53.4) | 1686 (59.5) | 1746 (48.4) | 407 (54.5) | <0.001 |
| Race/ethnicity |  |  |  |  | <0.001 |
| Non-Hispanic White | 3511 (48.8) | 1414 (49.9) | 1759 (48.7) | 338 (45.2) |  |
| Non-Hispanic Black | 1693 (23.5) | 593 (20.9) | 879 (24.4) | 221 (29.6) |  |
| Mexican American | 1691 (23.5) | 685 (24.2) | 843 (23.4) | 163 (21.8) |  |
| Other race/ethnicity | 296 (4.1) | 143 (5.0) | 128 (3.5) | 25 (3.3) |  |
| Education, years | 10.9 ± 3.8 | 11.4 ± 3.8 | 10.8 ± 3.8 | 9.8 ± 3.9 | <0.001 |
| Systolic BP, mmHg | 132.3 ± 19.6 | 115.5 ± 8.6 | 137.5 ± 11.1 | 170.2 ± 13.5 | <0.001 |
| Diastolic BP, mmHg | 76.4 ± 10.2 | 70.2 ± 6.1 | 79.5 ± 9.1 | 85.4 ± 13.8 | <0.001 |
| BMI, kg/m<sup>2</sup> | 27.7 ± 5.5 | 26.6 ± 5.2 | 28.4 ± 5.6 | 28.0 ± 5.6 | <0.001 |
| Diabetes mellitus | 761 (10.6) | 203 (7.2) | 433 (12.0) | 125 (16.7) | <0.001 |
| Smoking status |  |  |  |  | <0.001 |
| Never smoker | 3291 (45.8) | 1263 (44.6) | 1627 (45.1) | 401 (53.7) |  |
| Former smoker | 2255 (31.4) | 812 (28.6) | 1234 (34.2) | 209 (28.0) |  |
| Current smoker | 1645 (22.9) | 760 (26.8) | 748 (20.7) | 137 (18.3) |  |
| Total cholesterol, mg/dL | 217.3 ± 43.7 | 212.2 ± 43.4 | 219.9 ± 43.4 | 224.6 ± 44.0 | <0.001 |
| HDL cholesterol, mg/dL | 51.3 ± 16.3 | 52.1 ± 15.7 | 50.5 ± 16.6 | 52.4 ± 17.1 | <0.001 |
| BP lowering medication use | 1568 (21.8) | 261 (9.2) | 979 (27.1) | 328 (43.9) | <0.001 |
| Prolonged P-wave duration | 1828 (25.4) | 583 (20.6) | 1002 (27.8) | 243 (32.5) | <0.001 |
| Abnormal P-wave axis | 1686 (23.4) | 746 (26.3) | 773 (21.4) | 167 (22.4) | <0.001 |
| Abnormal DTNPV1 | 166 (2.3) | 45 (1.6) | 93 (2.6) | 28 (3.7) | <0.001 |
| AC burden |  |  |  |  | 0.003 |
| 0 markers | 4005 (55.7) | 1649 (58.2) | 1987 (55.1) | 369 (49.4) |  |
| 1 marker | 2713 (37.7) | 1007 (35.5) | 1386 (38.4) | 320 (42.8) |  |
| 2 markers | 452 (6.3) | 170 (6.0) | 226 (6.3) | 56 (7.5) |  |
| 3 markers | 21 (0.3) | 9 (0.3) | 10 (0.3) | 2 (0.3) |  |
| AC presence (≥1 marker) | 3186 (44.3) | 1186 (41.8) | 1622 (44.9) | 378 (50.6) | <0.001 |

DTNPV1, Deep terminal negativity in V1; AC, atrial cardiopathy; BP, blood pressure; HTN, hypertension; BMI, body mass index; HDL-cholesterol, high-density lipoprotein cholesterol
\*mean ± SD for continuous variables and frequency % for categorical variables

Over a median follow-up of 13.8 years, 183 stroke deaths occurred (overall rate 1.98 per 1,000 person-years). During follow-up, stroke mortality increased progressively with greater atrial cardiopathy (AC) burden. Stroke mortality rates rose from 1.31 per 1,000 person-years among participants with no AC markers to 2.51, 5.24, and 4.64 per 1,000 person-years among those with 1, 2, and 3 markers, respectively, although the estimate for participants with 3 markers was based on only one event. Stroke mortality also increased across blood pressure (BP) categories, with incidence rates of 1.01, 2.23, and 5.23 per 1,000 person-years among participants with normal/elevated BP, stage 1–2 hypertension, and severe hypertension, respectively. Kaplan–Meier curves showed progressively higher cumulative stroke mortality across combinations of increasing BP severity and the presence of AC (**Figure 1**). Participants with severe hypertension and AC had the highest cumulative stroke mortality, whereas those with normal/elevated BP and no AC had the lowest risk. Separation of the curves became more pronounced over time, supporting a joint effect of AC and BP on stroke mortality.

**Figure 1:**
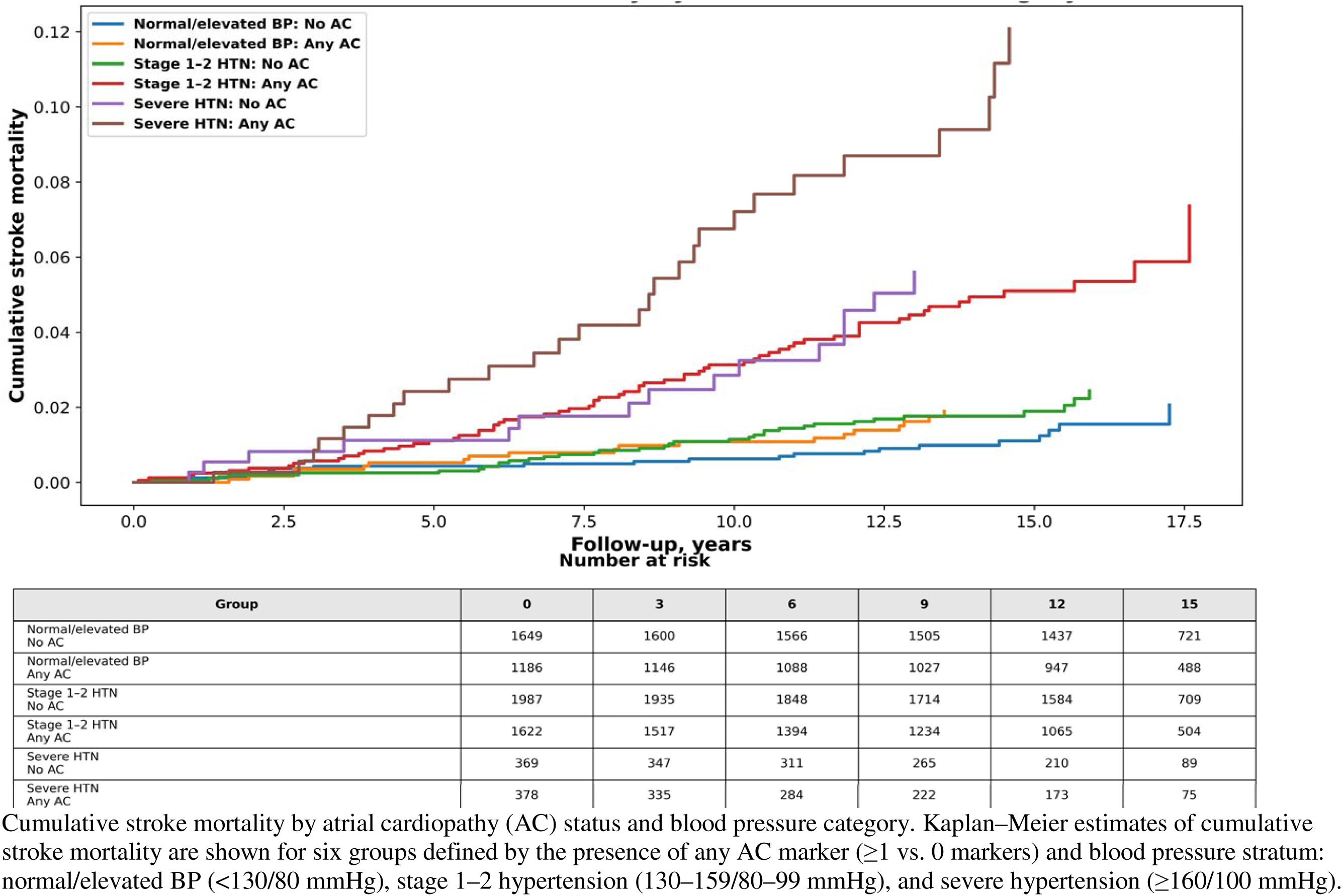
Cumulative stroke mortality by AC status and BP category.

In the overall cohort, each additional atrial cardiopathy (AC) marker was independently associated with a higher risk of stroke mortality after full adjustment (hazard ratio [HR], 1.41; 95% confidence interval [CI], 1.13–1.77; P=0.003). Similarly, the presence of any AC marker (≥1 vs. 0) was associated with a 49% higher risk of stroke mortality (HR, 1.49; 95% CI, 1.08– 2.05; P=0.016). The association between AC and stroke mortality was significantly modified by blood pressure (BP) category (interaction P=0.003 for AC burden). In BP-stratified analyses, the association between AC burden and stroke mortality strengthened progressively with increasing BP severity, with adjusted HRs of 0.88 (95% CI, 0.52–1.49) in participants with normal/elevated BP, 1.39 (95% CI, 1.03–1.88) in those with stage 1–2 hypertension, and 2.94 (95% CI, 1.82– 4.75) in those with severe hypertension. A similar pattern was observed for AC presence (≥1 vs. 0), with no significant association in the normal/elevated BP group (HR, 0.94; 95% CI, 0.48– 1.86), a borderline association in stage 1–2 hypertension (HR, 1.54; 95% CI, 0.99–2.40), and a significant association in severe hypertension (HR, 2.60; 95% CI, 1.27–5.32) (**Table 2**). However, the interaction between AC presence and BP categories for stroke mortality did not reach statistical significance (p=0.109).

**Table 2:** Association of Atrial Cardiopathy with Stroke Mortality Across Blood Pressure Categories.

|  | Atrial Cardiopathy status | HR (95% CI) |  |
| --- | --- | --- | --- |
|  |  | Model 1* | Model 2† |
| 0 RFs | AC Absent | 0.91 (0.48–1.72) | 0.94 (0.48–1.86) |
|  | AC Present | 0.89 (0.54–1.49) | 0.88 (0.52–1.49) |
| 1 RFs | AC Absent | 1.66 (1.08–2.54) | 1.54 (0.99–2.40) |
|  | AC Present | 1.45 (1.09–1.93) | 1.39 (1.03–1.88) |
| ≥ 2 RFs | AC Absent | 2.54 (1.27–5.09) | 2.60 (1.27–5.32) |
|  | AC Present | 2.89 (1.78–4.68) | 2.94 (1.82–4.75) |

Abbreviations: BP, blood pressure; HTN, hypertension; PY, person-years; HR, hazard ratio; CI, confidence interval.
\* Socio-demographic model adjusted for age, sex, race/ethnicity, and education.
† Fully adjusted model additionally adjusted for diabetes, smoking status, body mass index, total cholesterol, HDL cholesterol, and antihypertensive medication use. Interaction P=0.003 for AC burden, and 0.109 for AC present

## Discussion

In a US cohort of adults free of cardiovascular disease, ECG-defined atrial cardiopathy (AC) burden was independently associated with stroke mortality, with each additional AC marker conferring a 41% higher risk after multivariable adjustment. More importantly, this association was significantly modified by blood pressure (BP), strengthening progressively across increasing BP categories. Whereas AC burden was not significantly associated with stroke mortality among individuals with normal/elevated BP, the association became progressively stronger in those with stage 1–2 hypertension and was greatest among participants with severe hypertension, in whom each additional AC marker was associated with nearly a threefold higher risk of stroke mortality. Although similar trends were observed when AC was analyzed as a binary variable (AC presence ≥1 vs. 0 markers), the interaction with BP was no longer statistically significant, suggesting that cumulative AC burden may provide a more informative measure of atrial cardiopathy for risk stratification and future investigations of stroke risk.

Previous studies have shown that individual ECG markers of AC are associated with incident atrial fibrillation, stroke, heart failure, and mortality,^3,6,12,20^ and that increasing AC burden confers progressively greater risk.^10^ By demonstrating similar dose-dependent associations with stroke mortality, our study further supports the prognostic value of ECG-defined AC as a marker of adverse cerebrovascular risk.

The main novel finding of our study is that BP significantly modified the association between AC burden and stroke mortality. The significant interaction between AC burden and BP category (P for interaction = 0.003) indicates that the association between AC burden and stroke mortality strengthened progressively with increasing BP severity. Rather than acting as an isolated risk factor, the prognostic significance of AC appeared to depend on the underlying vascular milieu. Hypertension promotes structural and electrophysiological remodeling of the left atrium, including myocyte hypertrophy, interstitial fibrosis, atrial enlargement, and impaired mechanical function, all of which favor thrombus formation.^21^ In parallel, hypertension is associated with endothelial dysfunction, chronic inflammation, and altered hemodynamics that further enhance thrombotic susceptibility.^11,12,22^ The graded interaction observed in our study is therefore biologically plausible and suggests that progressive hypertensive vascular disease amplifies the thromboembolic potential of an already abnormal atrial substrate.

Our findings also emphasize the importance of considering hypertension as an effect modifier rather than solely as a confounder. Most prior studies investigating AC have adjusted for hypertension in multivariable models but have not examined whether the association between AC and stroke varies across BP levels.^2,3,8,12^ Our results indicate that stratification by BP reveals clinically meaningful heterogeneity that would otherwise remain obscured. The absence of a statistically significant interaction for binary AC (p=0.109) suggests the concept that cumulative burden of atrial abnormalities more accurately reflects disease severity and provides greater prognostic discrimination than dichotomous classification alone.

These findings may also help contextualize recent clinical trials evaluating AC as a therapeutic target. The ARCADIA trial failed to demonstrate superiority of apixaban over aspirin among patients with cryptogenic stroke and AC.^23^ One potential explanation is that the prognostic relevance of AC varies according to the underlying vascular phenotype. Recent exploratory analyses from ARCADIA showed that hypertension with high-risk features significantly modified the treatment effect of apixaban versus aspirin, with anticoagulation benefit confined to patients without high-risk HTN features.^24^ Although these observations differ from our findings at first glance, they may reflect complementary mechanisms. Severe hypertension may increase the prognostic importance of AC while simultaneously introducing competing non-cardioembolic stroke pathways that are unlikely to be prevented by anticoagulation alone.

Our findings have several potential clinical implications. Because ECG-derived P-wave markers are inexpensive, widely available, and routinely obtained in clinical practice, integrating AC burden with BP severity may improve identification of individuals at particularly high risk of stroke mortality. Individuals with both severe hypertension and AC may warrant closer surveillance and more intensive risk-factor modification, particularly aggressive BP control. Whether this subgroup derives differential benefit from anticoagulation or other targeted therapies remains uncertain and warrants prospective investigation.

The strengths of this study include the large, community-based racially diverse cohort, standardized centralized ECG processing, long-term mortality follow-up, and evaluation of AC using both cumulative burden and binary definitions. The consistency of findings across these alternative definitions further supports the robustness of the observed associations.

Several limitations merit consideration. First, ECG-defined AC may not fully capture structural and functional atrial abnormalities that can be detected by cardiac imaging. Second, residual confounding cannot be excluded despite comprehensive adjustment. Third, the outcome was limited to stroke mortality and therefore does not capture nonfatal cerebrovascular events. Finally, the observational design precludes causal inference.

### Conclusions

In this large US population-based cohort of adults free of cardiovascular disease, ECG-defined atrial cardiopathy burden was independently associated with stroke mortality, and its prognostic significance increased progressively with blood pressure severity. These findings identify hypertension as an important modifier of the association between atrial cardiopathy and stroke mortality and suggest that cumulative AC burden may provide a more informative measure than a binary definition for cerebrovascular risk stratification.

## Data Availability

The data used in this analysis are publicly available through the National Center for Health Statistics (NCHS) as part of the Third National Health and Nutrition Examination Survey (NHANES III) and can be accessed at https://wwwn.cdc.gov/nchs/nhanes/nhanes3/.

